# Temporal variation in one-time partnership rates among young men who have sex with men and transgender women

**DOI:** 10.1101/2020.10.19.20215178

**Authors:** Patrick Janulis, Steven M. Goodreau, Michelle Birkett, Gregory Phillips, Martina Morris, Brian Mustanski, Samuel M. Jenness

## Abstract

**Background:** Volatility in sexual contact rates has been recognized as an important factor influencing HIV transmission dynamics. One-time partnerships may be particularly important given the potential to quickly accumulate large number of contacts. Yet, empirical data documenting individual variation in contact rates remains rare. This study provides much needed data on temporal variation in one-time partners to better understand behavioral dynamics and improve the accuracy of transmission models.

**Methods:** Data for this study from a longitudinal cohort study of young men who have sex with men (MSM) and transgender women in Chicago. Participants provided sexual network data every 6-months for 2 years. A series of random effects models examined variation in one-time partnership rates and disaggregated within and between associations of exposure variables.

Exposure variables included prior number of one-time partners, number of casual partners, and having a main partner.

**Results:** Results indicated substantial between and within person variation in one-time partners. Casual partnerships were positively associated and main partnerships negatively association with one-time partnership rates. There remained a small positive association between prior one-time partnerships and the current number of one-time partnerships.

**Conclusions:** Despite the preponderance of a low number of one-time partners, substantial variation in one-time partnership rates exists among young MSM and transgender women. Accordingly, focusing on high contact rate individuals alone may be insufficient to identify periods of highest risk. Future studies should utilize these estimates to more accurately model how volatility impacts HIV transmission and better understand how this variation influences intervention effectiveness.

## INTRODUCTION

Men who have sex with men (MSM) continue to account for the majority of new HIV infections in the United States (Matthews et al., 2016), despite making up less than 5% of the population and the availability of highly effective HIV treatment and prevention tools (Sidibé et al., 2016). Current federal initiatives for ending the HIV epidemic (Fauci et al., 2019) prioritize efforts to identify, predict, and prevent new HIV infections among key populations such as young MSM, Black MSM, and transgender women who continue to experience extremely high HIV incidence (Crepaz et al., 2019). Yet, our ability to reduce HIV incidence in these populations relies on high-quality data to understand transmission risk and to design and deploy preventive interventions (Delva et al., 2016; Pellis et al., 2015).

Rates of sexual partnership acquisition have proven critical for understanding the spread of HIV and other sexually transmitted infections (STIs). While between-person heterogeneity in rates have been extensively studied among MSM (Millett et al., 2007, 2012; Rosenberg et al., 2011), a smaller number of studies have examined temporal variation in partner contact rates. For example, seasonality in contact rates has been documented and may be influenced by health (e.g., STI diagnoses) or personal factors (Carlo Hojilla et al., 2016), specific events such as travel (Elsesser et al., 2016), and prevention behaviors such as initiating pre-exposure prophylaxis (PrEP; Liu et al., 2013; Montaño et al., 2019). There is also evidence of variation in contact rates over longer periods of time (Lim et al., 2012; Romero-Severson et al., 2015) that may reflect both episodic change as well as developmental and historical trends (Basten et al., 2018; Swann et al., 2019). Given evidence documenting volatility in sexual contact rates, it is also important to understand the impact of this variation on HIV transmission risk.

Epidemic models examining this question have found within-person variation in partnership rates increases the population prevalence of HIV (Rozhnova et al., 2016) and increases the number of acute-stage HIV transmissions (Romero-Severson et al., 2013). These differences may be due to a higher likelihood that individuals are infected during high contact rate periods because individuals transitioning from low to high contact rates are more likely to be susceptible (Joseph Davey et al., 2017; X. Zhang et al., 2012). Dynamic variation in risk may also decrease the effectiveness of ‘test-and-treat’ strategies for HIV elimination (Rozhnova et al., 2016) but increase the effectiveness of PrEP (Rozhnova et al., 2019). Yet, many of these studies rely on behavioral data from earlier phases of the HIV epidemic (Romero-Severson et al., 2012, 2015) or that do not represent key populations to the current US HIV epidemic (Rozhnova et al., 2016, 2019) such as Black MSM and young MSM. Accordingly, our understanding of the impact of temporal variation in sexual contact rates among priority populations in the US remains limited.

This study aims to estimate within and between-person variation in one-time partnership contact rates using data from a large longitudinal cohort of young MSM and transgender women from Chicago. We focus our analysis on one-time partners because the short duration of one-time partnerships leads to a strongly right skewed distribution (i.e., a small number of individuals with a large number of one-time partners). Accordingly, longitudinal data on one-time partnership rates are vitally important so we can accurately understand stability or volatility in contact rates, including among the small number of high contact rate individuals who may have outsized impact on the spread of HIV. This information will be useful in both clinical settings in which risk assessment is conducted and for parameterizing epidemic transmission models that depend on these data as inputs.

## METHODS

### Participants and Procedures

Data for this analysis come from the RADAR study (Mustanski et al., 2019), an accelerated longitudinal (Duncan et al., 1996) cohort study of young MSM and transgender women. All participants in the RADAR study were required to meet the following criteria: 16 to 29 years old at enrollment, male assigned at birth (transgender women and other non-cisgender identified individuals were eligible), English-speaking, reported a sexual encounter with a man in the previous year or identified as gay or bisexual, and able to attend in-person research visits in Chicago. Recruitment for this study came from two prior cohorts of sexual and gender minority adolescents and young adults, Project Q2 (Mustanski et al., 2010) and Crew 450 (Mustanski et al., 2013), enriched with new cohort members recruited through venue-based, online, and peer recruitment. In addition, significant partners of RADAR cohort members were also eligible to enroll in the study. Expanded details regarding recruitment have been previously described (Mustanski et al., 2019). RADAR protocol was for enrolled participants to complete one study visit every six months for five years. Data for the current analysis comes from the beginning of data collection (February 2015) through February 2020. At the time of analysis, there were 1,053 active cohort participants enrolled in the RADAR study. However, we limited our analysis to participants who had completed their first six study visits using data from the second through the sixth visit, representing roughly 2 years of follow-up starting six months after enrollment. There were 944 participants eligible to have completed their sixth visit with 804 completing the 6th visit, indicating a completion rate of 85.2%.

### Measures

At each study visit, participants completed an inventory of their sexual, drug use, and social networks, a psycho-social survey, and biomedical specimen collection (HIV, STI, and drug screening). Demographic information (age, race/ethnicity, gender identity) was collected during a computer assisted self-interview, while sexual partnership data were collected via an interviewer-assisted network inventory. Detailed information regarding sexual partners was collected, including: indication of serious relationship, date of first and last sex (within the past six months), number of anal sex acts, and number of anal sex acts without a condom.

For the current analysis, we classified partnerships into three categories, as is typical for other MSM partnership studies (Sullivan et al., 2014; Weiss et al., 2019): main (i.e., serious), casual, and one-time partners. Main partners were those with whom the participant indicated they were “currently in a serious relationship.” Participants were asked to interpret ‘serious relationship’ however they would like, although they were also told, “This could be someone you call a boyfriend or girlfriend, significant other, partner, or husband or wife.” Casual partners were partners with whom the participant indicated having sex on more than a single day, but were not indicated as a main partner. One-time partners were partners whom the participant indicated the same date of first and last sex. For casual and one-time partners, partner counts reflect the cumulative number of partners in the six months prior to the study visit.

We also include time-fixed and time-varying exposures variables in our analysis. The time-fixed exposures variables were race/ethnicity and gender identity. Time-varying exposure variables included age, the number of current casual partners, the existence of a current main partner, number of casual partners during the previous observation, and existence of a main partners during the previous observation, and the number of one-time partners during the previous observation (i.e., autocorrelation).

### Analysis

In our preliminary analysis, we examined the extent of variation in one-time partnership throughout the observed study period using descriptive statistics and visualizations. First, we examined the number of participants in 4 overlapping groups based on their maximum or minimum number of one-time partners reported throughout the entire study, Second, we plotted the counts of one-time partners reported during a single visit, the number of one-time partners by age, and the number of one-time partners across study visits (i.e., wave).

In our primary analysis, we used a series of random effects models to examine correlates of one-time partnership using a negative binomial distribution. Specifically, we utilized a ‘hybrid’ random effects models (Falkenström et al., 2017; Firebaugh et al., 2013) to disaggregate within and between variation in our primary exposure variables (i.e., number of casual partners and having a main partner) using person-mean centering to capture within-person variation in these exposure variables. In addition, we added a person-mean exposure variable for casual and main partners to estimate the between-person effects on one-time partners.

Modeling took place in 3 stages. First, an intercept only model was examined using only a random intercept to estimate the extent to which one-time partnership rates vary within and between individuals. An intraclass correlation coefficient (ICC) was estimated using appropriate adjustments for the negative binomial distribution (Nakagawa et al., 2017). Next, a model including all exposure variables except time-lagged exposure variables was examined. Finally, a model with all previous variables and time-lagged exposure variables was examined including lagged variables for casual, serious, and one-time partners. There are well-known challenges with estimating models with lagged dependent variables because of the intrinsic correlation between the random intercept and the person-time error term (i.e., endogeneity) which violates a major assumption of linear models (Falkenström et al., 2017; Wilkins, 2018). Violating this assumption can cause problems such as substantially downward bias in estimates of other correlates (Achen, 2000). In order to examine for bias in our models due to endogeneity we: 1) first examined a model free of lagged terms so we can compare coefficients across models with and without a lagged dependent variable and 2) tested for autocorrelation in the residuals using a Breusch-Godfrey test, which has shown in simulations (Keele & Kelly, 2006) to be a robust indicator for detecting bias.

For all models, multiple imputation was utilized to account for 369 (9.2%) visits with any missing data. Specifically, we utilized multiple imputation by chained equations using predictive mean matching for count variables and logistic regression for dichotomous variables (Enders et al., 2016; van Buuren & Groothuis-Oudshoorn, 2010) with 20 imputed datasets. Aggregate model results from all 20 imputations are presented using averaged parameter estimates and standard errors (Rubin, 1987).

## RESULTS

Table 1 presents the demographics characteristics of participants (n = 804) and descriptive statistics regarding one-time partners across baseline and all study follow-up visits. The sample was predominantly Black/African-American (34.8%) or Hispanic (30.3%) with a mean age at baseline of 21.2 years (SD = 2.95). Additionally, this table includes descriptive statistics regarding categories of participants according to their number of one-time partners across the entire six waves of observation. The mean number of one-time partners in a single observation was 0.54 (SD = 1.22). Notably, a sizable minority of participants (42.1%) did not report any one-time partners during the entire study. Furthermore, a small (6.1%) number of participants reported at least a single one-time partner at each visit during the entire follow-up period.

**Table 1.** Baseline demographics and one-time partnership characteristics Demographics n (%)

| Demographics | n (%) |
| --- | --- |
| Race / Ethnicity |  |
| Black / African-American | 280 (34.8) |
| White | 189 (23.5) |
| Hispanic / Latino | 244 (30.3) |
| Other | 91 (11.3) |
| Gender |  |
| Male | 744 (92.5) |
| Transgender Female | 42 (5.2) |
| Other | 18 (2.2) |
| Age* | 21.2 (2.9) |
| One-time Partnerships |  |
| No One-Time Partners | 339 (42.1) |
| Never > 1 | 539 (67.0) |
| Never > 2 | 656 (81.5) |
| Never > 3 | 717 (89.2) |
| Never < 1 | 49 (6.1) |
| Never < 2 | 11 (1.4) |

Note. \*Age represents mean (SD)

Figure 1 shows the distribution of one-time partnerships during a single visit. As indicated, the most common number of partners during a 6-month period is zero with 72.6% of all visits in this category. Figures 2 and 3 use a random sample of 200 to display variation in one-time rates over age and wave. Figure 2 shows the trajectory of one-time partners across age with the mean number for each age group shown in blue. Figure 3 displays a heatmap of one-time partners across each observation wave, with each horizontal line representing a single participant. These figures demonstrate that, despite the overall low average number of one-time partners, participants do exhibit time-variation in the number of one-time partners.

**Figure 1.**
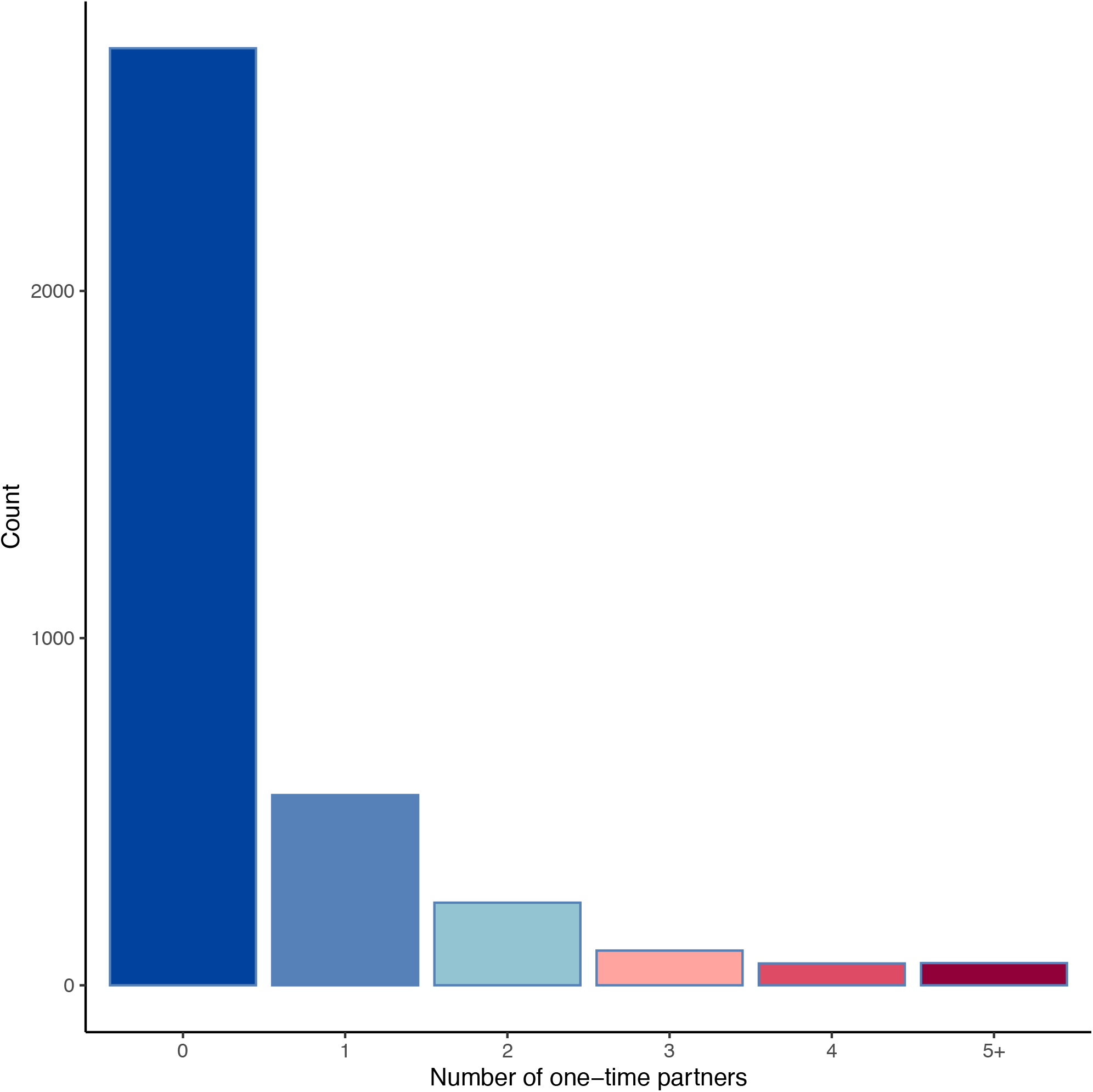
Number of one-time partners during a single visit.

**Figure 2.**
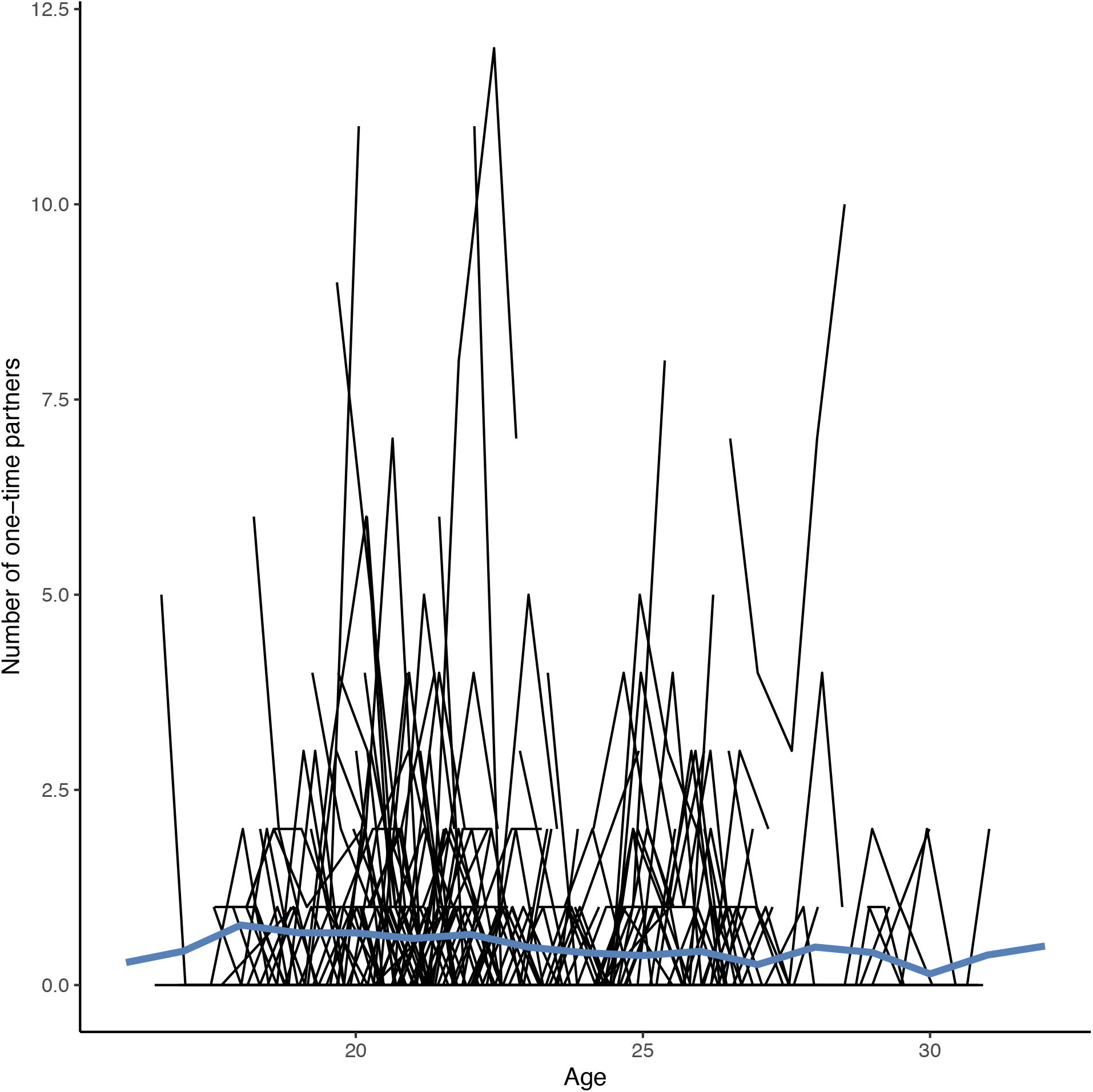
Number of one-time partners across age. Note. This figure represents data from a random sample of 200 participants for purposes for purposes of interpretability. The blue line represents the mean number of one-time partners across age.

**Figure 3.**
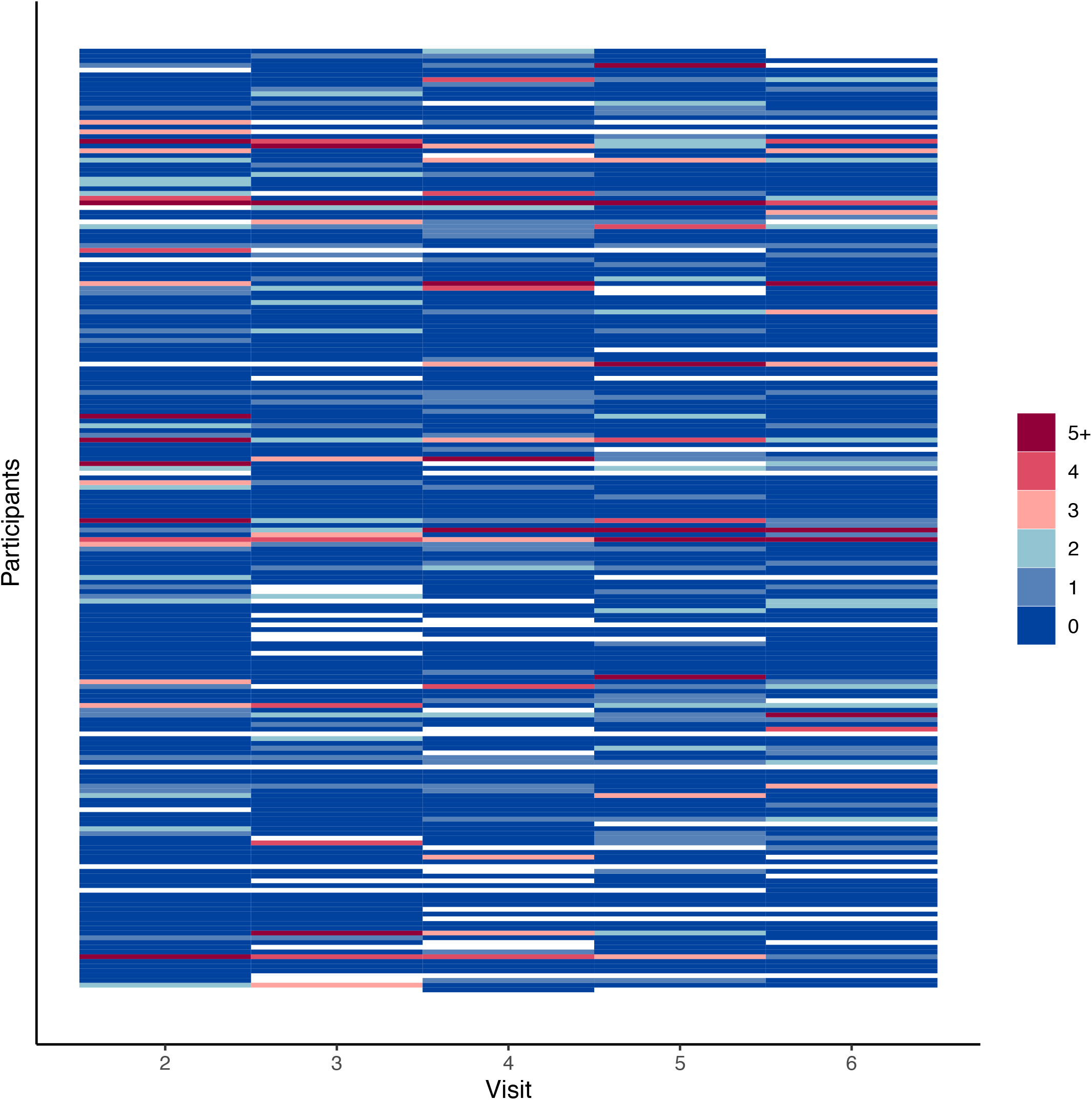
Number of one-time partners across visits. Note. This figure represents a data from a random sample of 200 participants. Horizontal changes in color across a single point on the y-axis represents temporal variation in the number of one-time partners.

The intercept-only model (Table 2) indicated that one-time partners varied both within and between individuals (ICC = 0.46), indicating 46% of the total variance in one-time partners occurred between individuals. The second model with exposure variables indicated several associations with one-time partners. Age was inversely associated with the number of one-time partners (IRR = 0.97 [95% CI: 0.94, 0.99]). Having more casual partners was associated with a higher number of one-time partners for both person-centered (IRR = 1.16 [95% CI: 1.08, 1.24]) and person-mean (IRR = 2.24 [95% CI: 1.91, 2.63]) exposure variables. In contrast, having a main partner was associated with having a lower number of one-time partners for both the person-centered (IRR = 0.50 [95% CI: 0.40, 0.63]) and person-mean (IRR = 0.49 [95% CI: 0.34, 0.69]) exposure variables. Finally, the third model with lagged exposure variables indicated that the previous number of casual partners was inversely associated with the number of one-time partners (IRR = 0.94 [95% CI: 0.89, 0.99]) and the prior number of one-time partners was positively associated with the current number of one-time partners (IRR = 1.09 [95% CI: 1.03, 1.15]) while all other associations remained the same direction with similar magnitude. The Breusch-Godfrey tests for the final models were not significant for any of the 20 imputed data sets (p-value range = 0.535 - 0.986) indicating these models did not have significantly autocorrelated residuals.

**Table 2.**
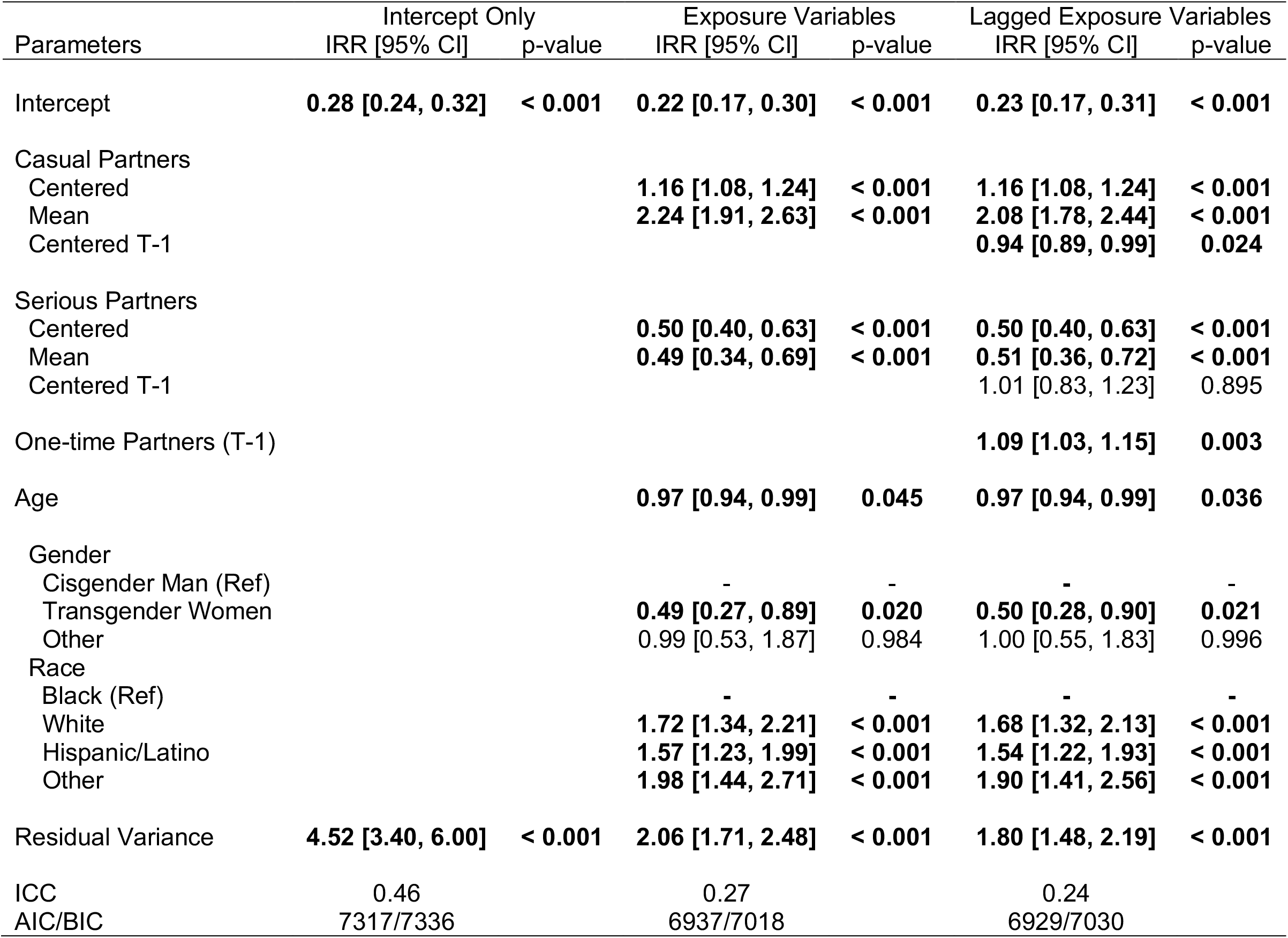
Results of random effects models of one-time partners.

| Parameters | Intercept Only |  | Exposure Variables |  | Lagged Exposure Variables |  |
| --- | --- | --- | --- | --- | --- | --- |
|  | IRR [95% CI] | p-value | IRR [95% CI] | p-value | IRR [95% CI] | p-value |
| Intercept | **0.28 [0.24, 0.32]** | **< 0.001** | **0.22 [0.17, 0.30]** | **< 0.001** | **0.23 [0.17, 0.31]** | **< 0.001** |
| Casual Partners |  |  |  |  |  |  |
| Centered |  |  | **1.16 [1.08, 1.24]** | **< 0.001** | **1.16 [1.08, 1.24]** | **< 0.001** |
| Mean |  |  | **2.24 [1.91, 2.63]** | **< 0.001** | **2.08 [1.78, 2.44]** | **< 0.001** |
| Centered T-1 |  |  |  |  | **0.94 [0.89, 0.99]** | **0.024** |
| Serious Partners |  |  |  |  |  |  |
| Centered |  |  | **0.50 [0.40, 0.63]** | **< 0.001** | **0.50 [0.40, 0.63]** | **< 0.001** |
| Mean |  |  | **0.49 [0.34, 0.69]** | **< 0.001** | **0.51 [0.36, 0.72]** | **< 0.001** |
| Centered T-1 |  |  |  |  | 1.01 [0.83, 1.23] | 0.895 |
| One-time Partners (T-1) |  |  |  |  | **1.09 [1.03, 1.15]** | **0.003** |
| Age |  |  | **0.97 [0.94, 0.99]** | **0.045** | **0.97 [0.94, 0.99]** | **0.036** |
| Gender |  |  |  |  |  |  |
| Cisgender Man (Ref) |  |  | - | - | - | - |
| Transgender Women |  |  | **0.49 [0.27, 0.89]** | **0.020** | **0.50 [0.28, 0.90]** | **0.021** |
| Other |  |  | 0.99 [0.53, 1.87] | 0.984 | 1.00 [0.55, 1.83] | 0.996 |
| Race |  |  |  |  |  |  |
| Black (Ref) |  |  | - | - | - | - |
| White |  |  | **1.72 [1.34, 2.21]** | **< 0.001** | **1.68 [1.32, 2.13]** | **< 0.001** |
| Hispanic/Latino |  |  | **1.57 [1.23, 1.99]** | **< 0.001** | **1.54 [1.22, 1.93]** | **< 0.001** |
| Other |  |  | **1.98 [1.44, 2.71]** | **< 0.001** | **1.90 [1.41, 2.56]** | **< 0.001** |
| Residual Variance | **4.52 [3.40, 6.00]** | **< 0.001** | **2.06 [1.71, 2.48]** | **< 0.001** | **1.80 [1.48, 2.19]** | **< 0.001** |
| ICC | 0.46 |  | 0.27 |  | 0.24 |  |
| AIC/BIC | 7317/7336 |  | 6937/7018 |  | 6929/7030 |  |

## DISCUSSION

In this study, we found that young MSM and transgender women reported both between and within person variation in one-time partners over time. Roughly half of the variance in one-time partners was accounted for by individual differences and half by within-person change. To the extent that one-time partnership rates help explain the spread of HIV and other STIs, these findings suggest focus on high-risk individuals alone will not fully capture the dynamic nature of this risk.

Descriptive statistics indicated that a large minority of participants (42.1%) never reported a one-time partner and participants reported no one-time partners during most visits (72.6%). These findings suggest that having a one-time partner is common, but varies over time in this target population. This finding could have important implications for the spread of HIV as individuals that have low contact rates are more likely to be susceptible when they transition to high contact rate periods (Joseph Davey et al., 2017; X. Zhang et al., 2012). While the focus of the current analysis on one-time partner contact rates makes direct comparisons to previous studies challenging since they have mainly examined aggregate risk scores (Basten et al., 2018; Pines et al., 2014; Wilkinson et al., 2017) or aggregated across all partner types (Lim et al., 2012; Romero-Severson et al., 2015), our main findings broadly concur with many of these studies that, while MSM vary in their contact rates and level of risk over time, there remains a preponderance of low-risk behavior. For example, one study (Wilkinson et al., 2017) found that a majority of participants were identified as ‘monogamous’ or ‘risk minimizers’ while another study found a majority of participants were ‘low risk’ in their trajectory (Pines et al., 2014), despite both studies indicating change in risk over time.

The current study also found that the previous number of one-time partners was weakly associated with the current number of one-time partners, indicating a small but statistically significant positive autocorrelation. This concurs with a previous study which also found a small positive autocorrelation in sexual contact rates among HIV-negative MSM (Romero-Severson et al., 2015). The small magnitude of this autocorrelation combined with substantial within-person variation and high prevalence of low contact rates with one-time partners provides important nuance to existing research on the ‘core’ group theory of STI transmission (Brunham & Others, 1997; Liljeros et al., 2003). This theory posits that a small number of individuals with high levels of turnover in their sexual contacts have a large impact on STI transmission. Although we did find a small group (6.1% of participants) that consistently reported at least a single one-time partner every six months, we also found a relatively modest level of autocorrelation and substantial within-person variation over time. Accordingly, these findings could suggest a ‘core group’ of individuals are unlikely to maintain a consistently high level of contact rates among one-time partners.

In addition, these results indicated casual and main partnerships likely play an important role in understanding variation in one-time partner contact rates. We found that having main partners, both within and between-person, was associated with having fewer one-time partners. This finding also concurs with previous studies indicating an inverse association between having a main partner and having non-main partners among MSM (Rosenberg et al., 2011) as would be expected given MSM report high rates of monogamous partnership agreements with main partners (Feinstein et al., 2018). In contrast, the number of casual partners was *positively* associated with the number of one-time partners, again for both within and between persons. The within person association (i.e., person centered) between casual partners and one-time partners supports the notion of ‘seasons’ of higher levels of sexual contact that are associated with health, psychological, and social contextual factors identified in a number of prior studies (Carlo Hojilla et al., 2016; Elsesser et al., 2016; Underhill et al., 2018). However, the between person association (i.e., person mean) suggests that individuals that tend to have higher numbers of casual partners on average also tend to report greater numbers of one-time partners. Accordingly, the highest rates of one-time partnership occur during periods of higher than average contact with casual partners (i.e., compared to an individual’s own average rate) amongst individuals that have higher levels of casual partners generally. Therefore, identifying additional correlates of these periods of high contact rates, in addition to the characteristics of the individuals that report above average levels of contacts (i.e., a core group), remains an important area for future research.

Together, these findings have important implications for understanding transmission dynamics for HIV in this target population. As noted, variation in risk behavior may substantially impact the spread of HIV and the effectiveness of interventions (Henry & Koopman, 2015; Romero-Severson et al., 2013, 2015; Rozhnova et al., 2016; X. Zhang et al., 2012). Previous studies have found temporal variation and trajectories may reduce the effectiveness of test and treat strategies (Rozhnova et al., 2016) while it could increase the effectiveness of PrEP (Rozhnova et al., 2019). Accordingly, the observed variation could impact the ideal combination of local HIV interventions (Nosyk et al., 2020). These findings also suggest that efforts to target high-risk individuals to prevent STIs (Giguère & Alary, 2015) may not yield the expected prevention benefits if these individuals do not maintain a consistent level of risk. However, to the extent that these temporary periods of high-risk can be understood and predicted, tailored interventions could be implemented for individuals during these periods. In fact, several studies have found that MSM report selectively utilizing preventions services such as PrEP during periods of high risk and report growing interest for ‘on-demand’ PrEP (Beymer et al., 2019; Cornelisse et al., 2019; Elsesser et al., 2016; Gafos et al., 2019; Saberi & Scott, 2020).

However, the detailed implications of these findings on the spread of HIV requires additional study. In particular, epidemic models that explicitly account for the observed variation are likely to provide the most accurate impact of these behaviors. For example, epidemic models are well positioned to estimate the impact of this temporal variation on intervention effectiveness (Hamilton et al., 2019; Luo et al., 2018), optimizing prevention policies (Jenness et al., 2016; Kelly et al., 2018), helping efficiently target resources (Elion et al., 2019; Goodreau et al., 2018), and estimating the cost-effectiveness of prevention strategies (Harmon et al., 2016; L. Zhang et al., 2019). Given the foundational work that has already examined the impact of episodic variation in contact rates on HIV (Henry & Koopman, 2015; Romero-Severson et al., 2015; X. Zhang et al., 2012), incorporating new parameters based on the current data to these models provides a promising future direction to contextual these findings and leverage them to inform public health action in this high priority population of young MSM and transgender women.

### Limitations

Limitations of the current analysis include the non-probability sample of RADAR participants that may not generalize to other populations of young MSM and transgender women. However, this large, recent, and racially/ethnically diverse sample of young MSM provides much needed data examining the extent of temporal variation in contact rates.

Furthermore, the focus of the current study on one-time partners makes it challenging to directly compare our findings with previous studies that have used broader (e.g., all partners) or specific (e.g., condomless sex acts) measures of HIV risk. Nonetheless, these partners remain important to understand given their strong right skew and limited existing data. The current study also utilized one modeling technique (i.e., random effects models) amongst a large number of potential modeling approaches to longitudinal data. It is possible other approaches such as observed Markov processes (Littman, 2009; Romero-Severson et al., 2015) may yield different insights.

### Conclusions

This study found within and between person variation in one-time partner contact rates among a large and diverse sample of young MSM and transgender women. Results indicated a small positive autocorrelation in one-time partnership rates, an inverse correlation with main partners, and a positive association with casual partners. Taken together this data provides much needed nuance to our understanding of temporal variation in partner contact rates in these priority populations. We hope future studies will continue to explore dynamic variation in partnership rates, how this variation impacts HIV/STI transmission, and leverage these insights to inform prevention activities.

## Data Availability

Original data is available through a data sharing agreement with the principal investigator of the RADAR study (Dr. Mustanski).

## Notes

FUNDING This work was supported by grants from the National Institute of Allergy and Infectious Diseases (R01AI138783) and the National Institutes on Drug Abuse (U01DA036939) at the National Institutes of Health.

CONFLICTS OF INTEREST The authors have no conflicts of interest to declare

### Competing Interest Statement

The authors have declared no competing interest.

### Funding Statement

This work was supported by grants from the National Institute of Allergy and Infectious Diseases (R01AI138783) and the National Institutes on Drug Abuse (U01DA036939) at the National
Institutes of Health.

### Author Declarations

This study was approved by the Northwestern University IRB

